# Will the Large-scale Vaccination Succeed in Containing the COVID-19 Epidemic and How Soon?

**DOI:** 10.1101/2021.04.16.21255543

**Authors:** Shilei Zhao, Tong Sha, Yongbiao Xue, Chung-I Wu, Hua Chen

## Abstract

The availability of vaccines provides a promising solution to containing the COVID-19 pandemic. Here, we develop an epidemiological model to quantitatively analyze and predict the epidemic dynamics of COVID-19 under vaccination. The model is applied to the daily released numbers of confirmed cases of Israel and United States of America to explore and predict the trend under vaccination based on their current epidemic status and intervention measures.

For Israel, of which 53.83% of the population was fully vaccinated, under the current intensity of NPIs and vaccination scheme, the pandemic is predicted to end between May 14, 2021 to May 16, 2021 depending on an immunity duration between 180 days and 365 days; Assuming no NPIs after March 24, 2021, the pandemic will ends later, between July 4, 2021 to August 26, 2021. For USA, if we assume the current vaccination rate (0.268% per day) and intensity of NPIs, the pandemic will end between February 3, 2022 and August 17, 2029 depending on an immunity duration between 180 days and 365 days. However, assuming an immunity duration of 180 days and with no NPIs, the pandemic will not end, and instead reach an equilibrium state with a proportion of the population remaining actively infected.

Overall the daily vaccination rate should be chosen according to the vaccine efficacy and the immunity duration to achieve herd immunity. In some situations, vaccination alone cannot stop the pandemic, and NPIs are necessary both to supplement vaccination and accelerate the end of the pandemic. Considering that vaccine efficacy and duration of immunity may be reduced for new mutant strains, it is necessary to remain cautiously optimistic about the prospect of the pandemic under vaccination.

---

Coronavirus disease 2019 (COVID-19) has rapidly spread to more than 200 countries and territories since the first cases reported in Wuhan, China on 31 December 2019, and the pandemic is still ongoing. As of March 24, 2021, COVID-19 has resulted in more than 131 million infections, causing around 2.85 million deaths. Given the high infectivity and limited clinically effective medicine for COIVD-19, a variety of non-pharmaceutical interventions (NPIs) were implemented world-wide intending to diminish the viral transmission and outbreaks, such as restrictions on movement, ban on the gatherings, social distancing, etc. The effectiveness of the NPIs in mitigating the viral spread have been reported in previous studies (*1–3*).

Vaccines become the crucial solution to containing the pandemic. Currently over 200 SARS-CoV-2 vaccine candidates are under swift development with different technology, including inactivated virus vaccines, live attenuated vaccines, recombinant protein vaccines, replication-incompetent vector vaccines, replication-competent vector vaccines, inactivated virus vector vaccines, DNA vaccines and RNA vaccines (*4*). Several candidates have completed the phase III human clinical trials and efficacy testing (*5*). The announced efficacy of vaccines is 95% for Pfizer/BioNTech (RNA vaccines) (*6*); 91.6% for Gamaleya (recombinant protein vaccines) (*7*); 94.1% for Moderna (RNA vaccines) (*8*); 70.4% for AstraZeneca (replication-incompetent vector vaccines) (*9*); 79.34% for Sinopharm (inactivated virus vaccines) (*10*); 50.65% *∼* 91% for Sinovac (inactivated virus vaccines) (*11*). The total capacity by 2021 is estimated to be around 7.4 billion doses (*12*), covering less than half of the global population (for most of COVID-19 vaccines 2 doses are required to get fully vaccinated). On December 14, 2020, UK became the first country starting the vaccination scheme, and as of March 24, 2021, more than 80 countries have started the vaccinations. Among these countries, Israel has the highest full vaccination rate of 53.83%, and USA has the largest vaccination doses of 130.47 million.

Although the availability of vaccines is a remarkable step towards the ending of COVID-19 pandemic. The prospect of the pandemic control is still unclear. It is important to quantitatively explore and predict the epidemic trend under the current speed of vaccination, vaccine efficacy and the strength of NPIs. And even more important, is to explore and evaluate the optimal scheme of vaccination. In this study, we develop an epidemic model to explore the pandemic dynamics under vaccination and NPIs. We apply the model to analyze and predict the pandemic trend of Israel and USA given their daily vaccination rates and other parameters, considering Israel is a leading country in vaccination and USA has a much larger population size and with the most severe COVID-19 pandemic. We also provide a formula for choosing speed of vaccination under different vaccine efficacy, immunity duration and NPIs. These results provide insights on efficient COVID-19 epidemic control under vaccination.

## Model

Based on our previous work SUQC (Suceptible -Unquarantined-Quarantined-Confirmed) (*3*), we develop a new model, named SUVQC (Suceptible-Unquarantined-Vaccined-Quarantined-Confirmed), to describe the epidemic dynamics of COVID-19 under vaccination. SUVQC includes nine variables corresponding to different states of an individual from the population (Fig. 1):

**Figure 1:**
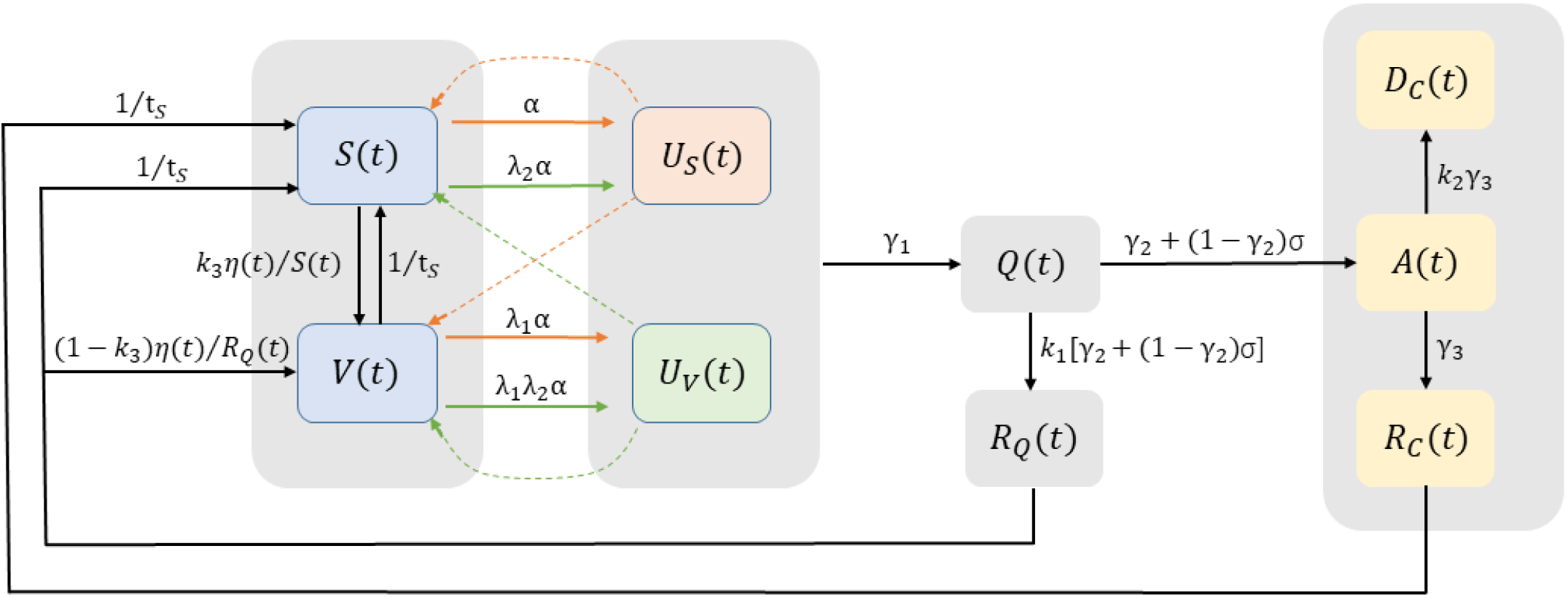
An schematic illustration of the model.

*S*(*t*), the number of susceptible individuals in the population at time *t*. For COVID-19, all individuals are assumed to be susceptible to the disease, except for those gain immunity through vaccination or infection. Note that the immunity against COVID-19 is not permanent, and an infected or vaccinated person becomes susceptible again when it is beyond the protection period of immunity.

*V* (*t*), the number of fully vaccinated individuals at time *t* that are still actively under protection with immunity gained through vaccination.

*U* (*t*), the number of infected individuals who are infectious and un-quarantined. *U* (*t*) includes two groups: *U*_*S*_(*t*), who are un-quarantined infected individuals from susceptible individuals, and *U*_*V*_ (*t*), who are from vaccinated individuals. *U* (*t*) = *U*_*S*_(*t*) + *U*_*V*_ (*t*). *U*_*V*_ (*t*) *>* 0 is due to the fact that COVID-19 vaccines cannot provide 100% protection.

*Q*(*t*), the number of quarantined infected individuals. The quarantined infected individuals have no contact with susceptible and vaccinated individuals and thus cause no infection in the population. Note that for simplicity, it also includes the infected individuals who have never been quarantined and finally transfer their state to *Q*(*t*) due to recovery and loss of infectivity.

*R*_*Q*_(*t*), the number of individuals among the quarantined, who were originally infected but non-confirmed, and are currently recovered with active immunity to the virus.

*A*(*t*), the number of confirmed infected cases who are still in active infection state, and are not recovered or dead yet.

*R*_*c*_(*t*), the number of recovered individuals who were confirmed cases and are still immune to the virus.

*D*_*c*_(*t*), the number of dead individuals among the confirmed infected cases.

Other than the above nine variables, we also define *C*(*t*), the cumulative confirmed infected cases. 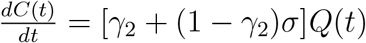.

The model includes 12 parameters:

*t*_*s*_, the average duration of immunological memory of individuals who recover from infection.

*t*_*v*_, the average duration of immunological memory of vaccinated individuals.

*α*, the infection rate, which is the average number of newly infected cases from *S* that is caused by an un-quarantined infected in *U*_*S*_ per day.

*η*(*t*), the daily number of individuals fully vaccinated.

*λ*_1_, the reduced probability for a vaccinated susceptible individual being infected. The **vaccine efficacy** is defined as 1 − *λ*_1_.

*λ*_2_, the reduced infectivity of vaccinated infected individual compared to *U*_*s*_, that is, the average infection rate of *U*_*v*_ is *λ*_2_*α*.

*γ*_1_, the quarantine rate for an un-quarantined infected being quarantined.

*γ*_2_, the confirmation rate of *Q*, is the probability that the quarantined infected are identified to be confirmatory cases by a conventional method.

*σ*, the subsequent confirmation rate of those infected that are not confirmed by the conventional methods, but confirmed with some additional tests.

*k*_1_, the ratio of un-confirmed infected cases over confirmed infected cases.

*k*_2_, the ratio of dead to recovered individuals among all confirmed cases.

*γ*_3_, the recovery rate of active confirmed infected individuals.

The transition among different states during the epidemic dynamics is summarized in Equation 1, and demonstrated in Fig. 1. The susceptible individuals *S* become *V* after vaccination at a constant rate. *S* can also move to *U*_*s*_ due to infection. Similarly, the vaccinated individuals *V* still have some chance of getting infected, and thus transit to *U*_*v*_. The infected individuals *U*_*s*_ and *U*_*v*_ move to *Q* by quarantine measures and lose the infectivity. Among individuals in *Q* state, some are confirmed, and later become recovered or dead. The rest individuals of *Q* are not confirmed by tests or other approaches, and move to *R*_*Q*_ directly. Note that the *Q* → *R*_*Q*_ flow may be due to the lack of test kits, or the infected cases with no symptoms (asymptomatic infection). It is reasonable to assume that all individuals without being confirmed are with mild symptoms, and thus no flow of *Q* → *D*_*Q*_ directly. Individuals from *V, R*_*c*_ and *R*_*Q*_ finally become susceptible to the disease again after a period of time, since the immunity against COVID-19 is likely not permanent. Since we are unable to distinguish between un-vaccinated individuals in *R*_*Q*_ and *S* at time *t, η*(*t*) includes two parts: *k*_3_*η*(*t*) individuals transiting from *S* to *V*, and (1 *− k*_3_)*η*(*t*) individuals transiting from *R*_*Q*_ to *V*, where 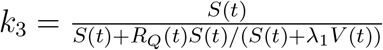.

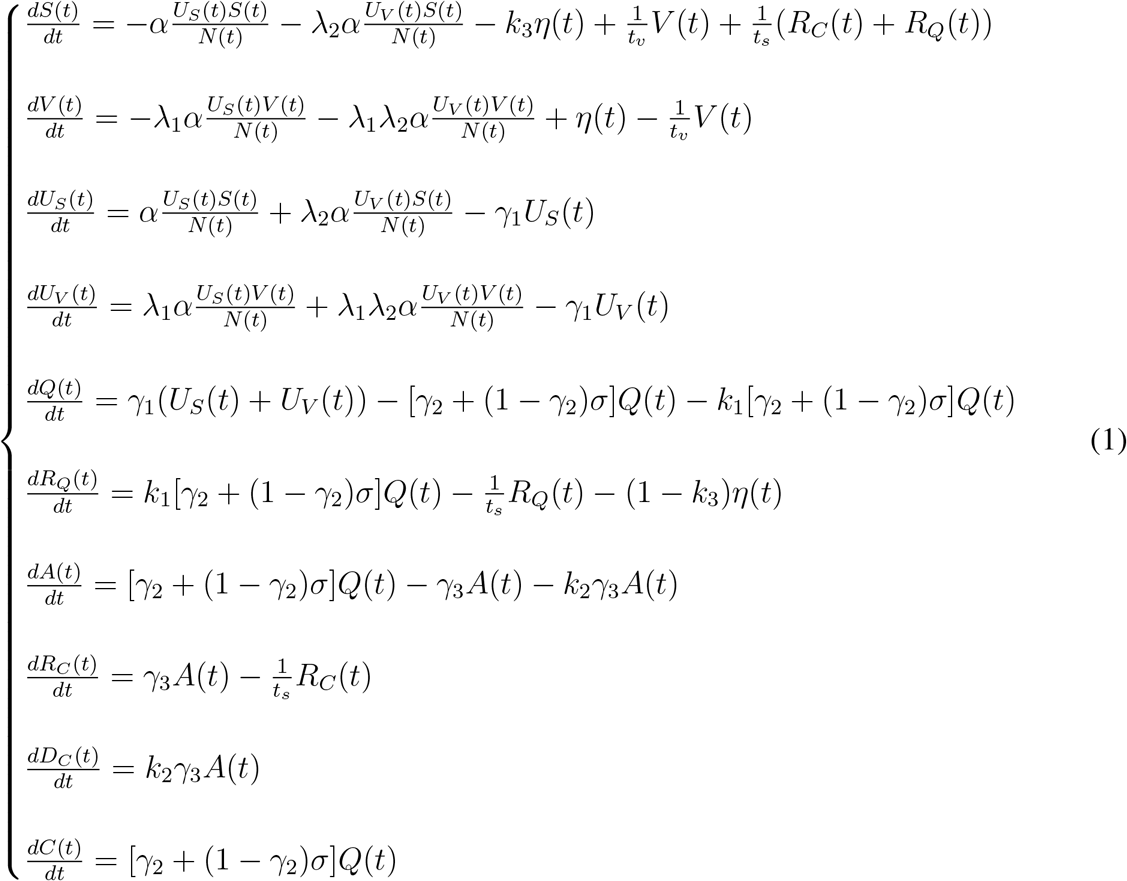

## Parameter Inference

The observed data we can use in the modeling fitting is the series of numbers of daily reported infected cases. Since the SUVQC model includes multiple parameters, for most of which, the data provides very limited information, we thus infer nine parameters and the initial values of eight variables using information from other sources, including known facts and previous studies:

- *t*_*s*_ and *t*_*v*_: A recent study demonstrated that the immune memory caused by SARS-CoV-2 infection can last up to 8 months after infection (*13*). Several other studies also suggested an immunization period of at least 6-9 months (*14–16*). At present, there is no clear conclusion on how long the immune protection induced by the vaccine can last. We set it to *t*_*s*_ = *t*_*v*_ = 240 days. Since the duration of immunity is critical to the epidemic dynamics, we also try three different durations of immunity, including 90 days, 180 days and 365 days.
- *α* = 0.2967: obtained by fitting an exponential curve to the numbers of confirmed cases in Wuhan in the very early phase of the pandemic (*3*).
- *λ*_1_ and *λ*_2_: the vaccines used so far in Israel and USA are Pfizer/BioNTech and Moderna, with the efficacy of 95% and 94.1% respectively (*6, 8*). Thus we set *λ*_1_ = 0.95. No epidemic data is available for estimating *λ*_2_ yet. It is reported that vaccines can reduce the incidence of severe illness, which may imply lower infectivity of *U*_*v*_ than *U*_*s*_. Nevertheless, even the asymptomatic patients have similar viral load to that in the symptomatic patients (*17*). Thus we assume *U*_*v*_ has the same infectivity as *U*_*s*_ and set *λ*_2_ = 1.
- *γ*_3_: the recovery rate of active confirmed cases is set to be *γ*_3_ = 1*/*14, since the mean hospital admission days for survivors and non-survivors are 11.3 days and 14.5 days respectively according to former studies (*18*).

The values of some other parameters and variables, e.g., *k*_1_, *k*_2_, *η, V*_0_, and *R*_*C*_(0) are population-specific, and will be elaborated in following sections.

*U*_*s*_(0), the initial number of un-quarantined infected individuals from susceptible individuals, *Q*(0), the initial number of quarantined infected individuals, and parameters *γ*_1_, *β* = *γ*_2_ + (1 *− γ*_2_)*σ* are inferred by fitting the numbers of daily reported cumulative confirmed cases *C*(*t*) with the SUVQC model. The loss function is defined as (*3*),

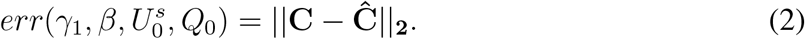

where 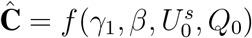 is the expected cumulative confirmed case numbers from the model (Eqn 1), which is solved by the fourth order Runge-Kutta method. The values of the four parameters or variables are inferred by optimizing the loss function with the interior-point method (implemented with fmincon in MATLAB).

## The epidemic trend of Israel under vaccination

The population size of Israel is around 9.05 million. The COVID-19 pandemic in Israel started on February 21, 2020. The country has undergone three waves of outbreaks (see https://en.wikipedia.org/wiki/COVID-19_pandemic_in_Israel). We inferred the temporal dynamics of *R*_*t*_ as a function of time by applying the SUQC model to daily reported confirmed cases using the sliding window scheme (with a window size of 3 weeks and a step size of one week; blue curve, fig. S1). It demonstrates the development and fluctuation of the pandemic during the process, being consistent with the public reports. The Israel government issued a series of control measures during the period, including social distancing, home isolation, travel bans, and restriction in movements etc. The Government Stringency Index provided by OxCGRT (*19, 20*) couples well with the *R*(*t*) trend, indicating the efficiency of multiple intervention measures (red curve, fig. S1). As of March 24, 2021, 830, 028 dynamics individuals have been reported to be infected by COVID-19. The Israel government procured COVID-19 vaccines from multiple sources: 8 million shots from Pfizer; 10 millions doses from AstraZeneca; and 6 million doses from Moderna. Vaccination has started since December 19, 2020, and is progressing steadily and rapidly. As of March 24, 2021, around 53.83% of the Israel population has been fully vaccinated. Together with other measures, vaccination demonstrates its efficiency, and the new confirmed cases demonstrated a persistent downward trend since Jan 20, 2021. We analyze the epidemic dynamics of COVID-19 in Israel under vaccination, and predict its future trend with the SUVQC model under parameter settings inferred from the reported data up to March 24, 2021. The prediction of future trend starts from March 24, 2021.

Some population-specific parameters and variables are inferred beforehand with prior knowledge:

1. The proportion of infected individuals remaining asymptomatic throughout infection is chosen to be 31% according to former studies (*21*), that is, *k*_1_ = 0.31*/*(1 *−* 0.31) = 0.45;
2. *η* = 54, 753 (0.605% of population), the number of daily vaccinated individuals is estimated as the average of numbers from 30 days (2021-2-23 to 2021-3-24) according to data from ourworldindata website (https://ourworldindata.org/). Note that *V* (*t*) is defined as the number of fully vaccinated individuals, and *η* is the daily number of individuals of fully vaccinated. This is a simplification of the reality since individuals not fully vaccinated are also immune to COVID-19 in a lower efficacy;
3. *V*_0_ = 3, 158, 669, the number of initial fully vaccinated individuals at Feb 23, 2021, is adopted from ourworldindata website (https://ourworldindata.org/).
4. *k*_2_ = 0.74%, the ratio of dead and recovered individuals among confirmed cases, *C*(0) = 759, 572, the initial confirmed cases, *D*_*C*_(0) = 5, 634, the initial confirmed deaths, and *A*(0) = 41, 610 are adopted from Johns Hopkins University Coronavirus Center (*22*).
5. *R*_*C*_(0) = 756, 439, the number of initial confirmed recovered individuals that still stain immunological memory to the virus is estimated by the recurrence formula *R*_*C*_(*t*) = (*C*(*t*) *− R*_*C*_(*t −* 1))/*t*_*s*_. *R*_*Q*_(0) is calculated by *R*_*Q*_(0) = *k*_1_*R*_*C*_(0).

Some other parameters, including *t*_*s*_ = 240 days, *t*_*v*_ = 240 days, *α* = 0.2967, *λ*_1_ = 0.05, *λ*_2_ = 1, *γ*_3_ = 1/14 are universal across different countries and chosen according to section “Parameter Inference”.

We set *U*_*V*_ (0) = 0, since *V* (*t*) *→ U*_*V*_ (*t*) is small compared to *S*(*t*) *→ U*_*S*_(*t*) at the early stage of the vaccination. The initial number of un-quarantined infected individuals from susceptible individuals *U*_*s*_(0), the initial number of quarantined infected individuals *Q*(0), and parameters *γ*_1_, *β* = *γ*_2_ + (1 *− γ*_2_)*σ* are inferred by fitting the numbers of daily reported cumulative confirmed cases *C*(*t*) with the SUVQC model. After obtaining all the values of parameters and initial variables, the initial value of susceptible individuals in the population *S*(0) is calculated with *S*(0) = *N − V* (0) *− U*_*S*_(0) *− U*_*V*_ (0) *− Q*(0) *− R*_*Q*_(0) *− A*(0) *− R*_*C*_(0) *− D*_*C*_(0). We explore the trends for two scenarios: one with NPIs, for which we assume the current control measures will continue with the same intensity; the other without NPIs, for which we assume that no more NPIs are imposed when under vaccination.

Fig. 2 presents the inference and prediction of epidemic dynamics in Israel assuming an eight-month duration of immunological memory. The SUVQC model is fitted to the daily numbers of confirmed cases from February 23, 2021 to March 24, 2021, and the unknown parameters and variables are inferred to be *U*_*s*_(0) = 38, 835, *Q*(0) = 26, 736, *γ*_1_ = 0.2714 and *β* = 0.0983. The model fits well with the observed data (Fig. 2A, red points, *R*^2^ = 0.9991). If we assume the intensity of NPIs remains as that of between February 23, 2021 and March 24, 2021 (the corresponding mean Government Stringency Index of 56.82 by OxCGRT. For comparison, the maximum Government Stringency Index of Israel is 94.44 around mid April, 2020, and 68.33 between March 1, 2020 and March 24, 2021), the epidemic in Israel is estimated to end around May 15, 2021 (with zero new confirmed case as the criterion), with 836, 059 cumulative confirmed infected cases and 6, 495 confirmed deaths. The number of active cases (*A*(*t*)) is estimated to reach peak at Mar 5, 2021, with the maximum value of *A*(*t*) = 46, 629 (Fig. 2, A and B).

**Figure 2:**
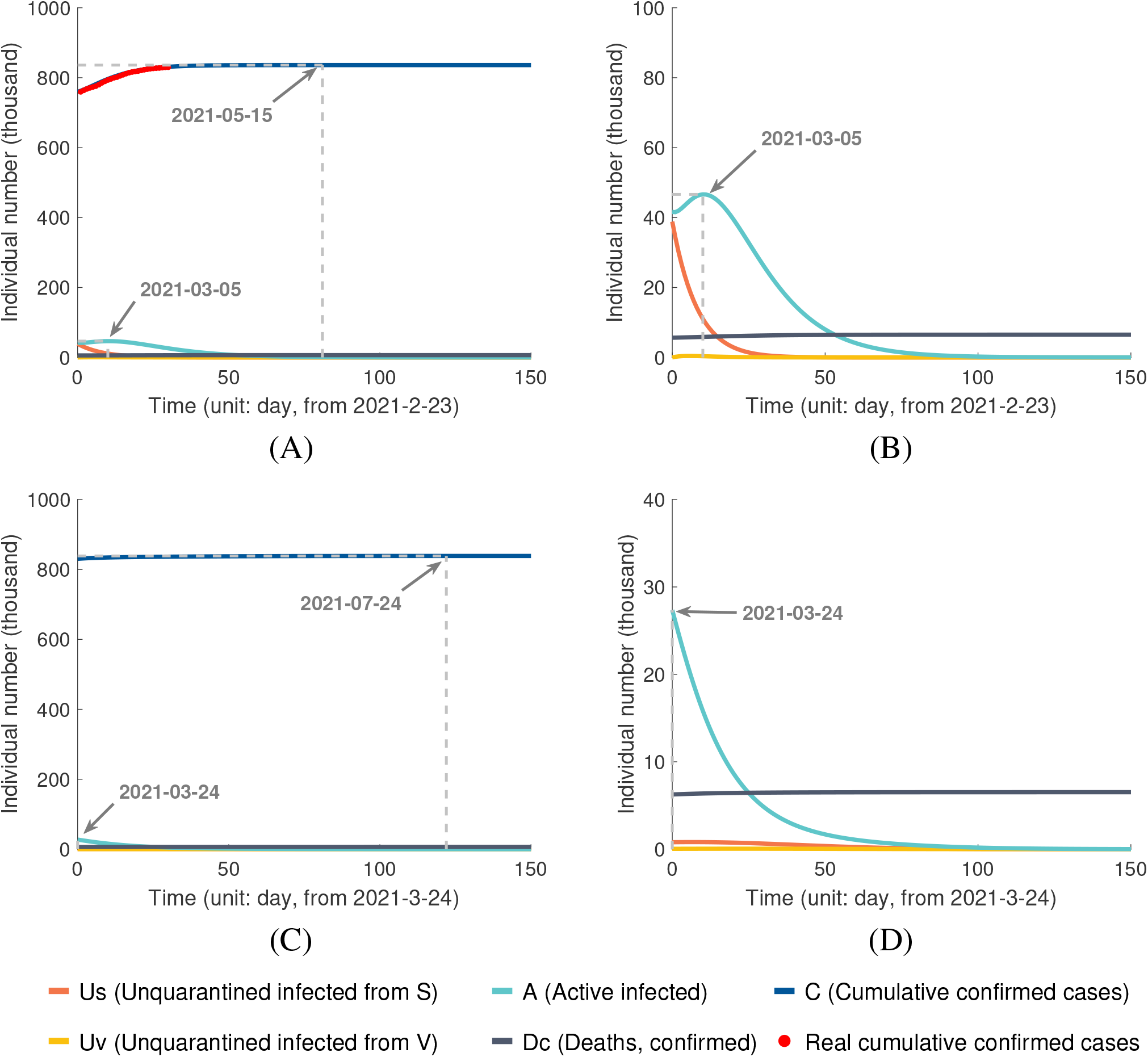
Inference and prediction of epidemic dynamics in Israel: (A) Parameter setting: vaccine efficacy 1 − *λ*_1_ = 0.95, daily vaccination rate *η/N* = 0.605% per day, duration of immunity *t*_*s*_ = 8 months, and with NPIs. (B) A zoom-in plot of (A) without *C*. (C) Parameter setting: all the parameters remain the same as (A) except that no NPIs after Mar 24 2021. (D) A zoom-in plot of (C) without *C*.

For the scenario assuming no more NPIs after March 24, 2021, the epidemic in Israel is estimated to end around July 24, 2021, with 838, 632 cumulative confirmed infected cases and 6, 520 confirmed deaths. The number of active cases is estimated to peak on March 24, 2021, with the value of *A*(*t*) = 27, 356 (Fig. 2, C and D).

To assess the impact of immunity duration of vaccines, we also carry out the same analysis with other possible *t*_*s*_ values, e.g., *t*_*s*_ = *t*_*v*_ = 90, 180, 365 days. The results are summarized in Table 1 (see the corresponding plots in Fig. S3-S5). Assume that maintaining the same intensity of NPIs as that of between February 23, 2021 and March 24, 2021, the epidemic in Israel will end between May 14, 2021 and May 19, 2021, depending on different durations of immunity. The number of cumulative confirmed infected cases is estimated to be between 835, 944 and 836, 576, with the number of deaths between 6, 493 and 6, 500. The epidemic in Israel is estimated to last for much longer time if no NPI after March 24, 2021, ranging from July 4, 2021 to August 26, 2021, with the number of cumulative confirmed infected cases between 837, 656 and 840, 383, and the number of deaths between 6, 513 and 6, 533. However, the epidemic in Israel will never end if the duration of immunity is only 90 days and with no NPIs. After several waves of fluctuations, the epidemic reaches an equilibrium state, with 68, 742 active infected cases (Fig. 2, C and D, Table 1). The vibrations are due to the initial proportions of vaccinated individuals and susceptible individuals. Overall, the epidemic trend of Israel under vaccination is optimistic if the duration of immunity is longer than 6 months.

**Table 1:**
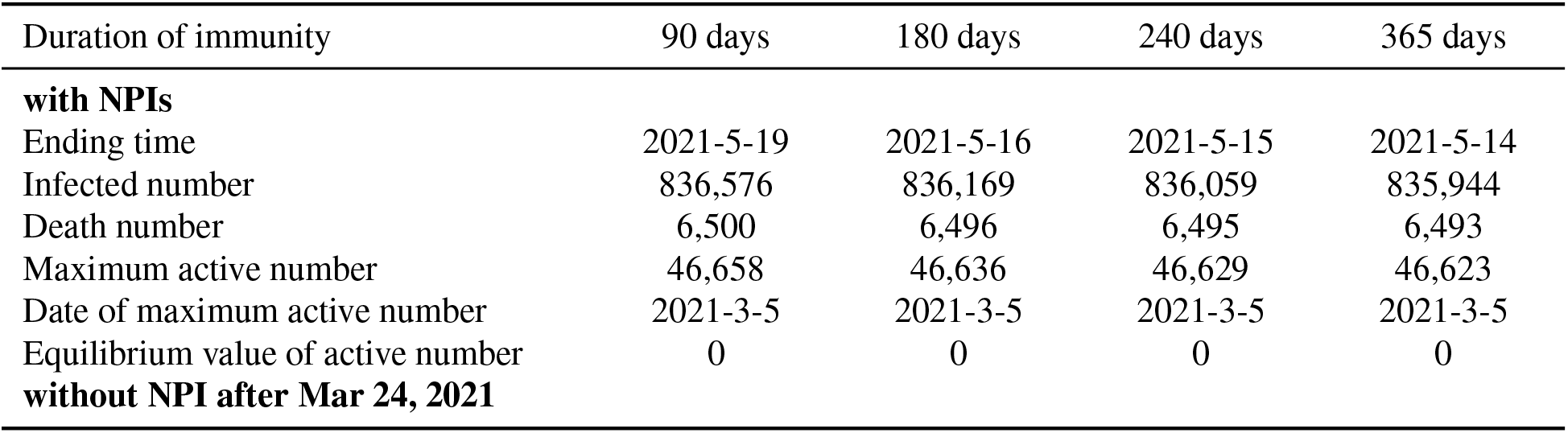
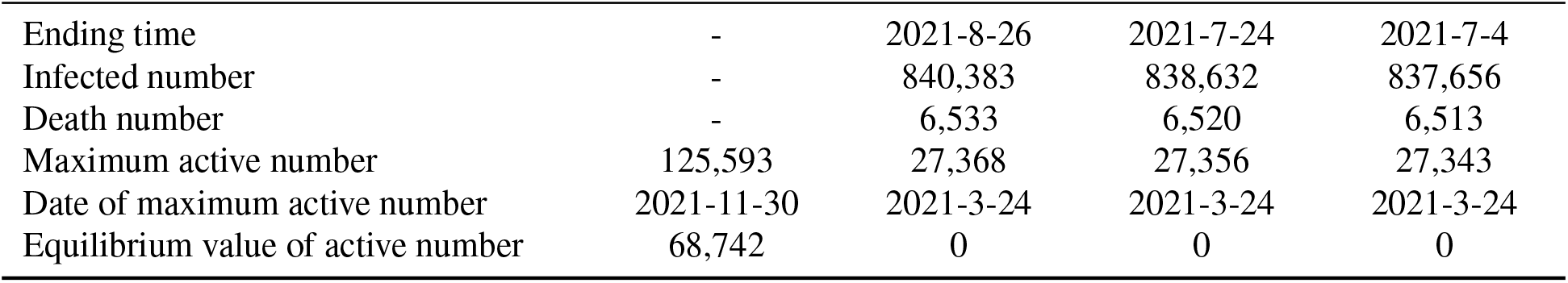
Prediction of the epidemic in Israel with different durations of immunity

| Duration of immunity | 90 days | 180 days | 240 days | 365 days |
| --- | --- | --- | --- | --- |
| **with NPIs** |  |  |  |  |
| Ending time | 2021-5-19 | 2021-5-16 | 2021-5-15 | 2021-5-14 |
| Infected number | 836,576 | 836,169 | 836,059 | 835,944 |
| Death number | 6,500 | 6,496 | 6,495 | 6,493 |
| Maximum active number | 46,658 | 46,636 | 46,629 | 46,623 |
| Date of maximum active number | 2021-3-5 | 2021-3-5 | 2021-3-5 | 2021-3-5 |
| Equilibrium value of active number | 0 | 0 | 0 | 0 |
| **without NPI after Mar 24, 2021** |  |  |  |  |
| Ending time | - | 2021-8-26 | 2021-7-24 | 2021-7-4 |
| Infected number | - | 840,383 | 838,632 | 837,656 |
| Death number | - | 6,533 | 6,520 | 6,513 |
| Maximum active number | 125,593 | 27,368 | 27,356 | 27,343 |
| Date of maximum active number | 2021-11-30 | 2021-3-24 | 2021-3-24 | 2021-3-24 |
| Equilibrium value of active number | 68,742 | 0 | 0 | 0 |

## The trend of epidemic in United States of America under vaccination

The population size of USA is around 327 million. The COVID-19 pandemic in US started around late January, 2020. The temporal dynamics of *R*(*t*) of USA is also inferred by applying the SUQC model to the daily reported confirmed cases with a sliding window scheme (with a window size of 3 weeks and a step size of one week; blue curve, Fig. S2). From the *R*(*t*) curve, we similarly identify three waves of outbreaks: the early stage before early April, from June 6 to August 8, and from early-October to late-December. The Government Stringency Index provided by OxCGRT (*19*) (*20*) couples well with the *R*(*t*) trend, which demonstrates the fluctuation of pandemic is highly correlated with the public events and implementation of controls by federal and state governments (red curve, fig. S2). As of March 24, 2021, 30, 012, 522 confirmed infected cases have been reported. The USA government ordered 600 million shots from Pfizer and Moderna, and before the end of July 2021, each company is expected to deliver 300 million doses in regular increments. Vaccination in USA has started since December 14, 2020. As of March 24, 2021, around 13.86% of the USA population have been fully vaccinated.

We also infer some parameters and variables before model-fitting with prior knowledge:

1. As of mid November 2020, 14.3% of the US population (around 47 millon) were estimated to be infected by SARS-CoV-2 (*23*), and at the same time, the reported confirmed cases is around 11 million. Thus *k*_1_ = (47 *−* 11)*/*11 = 3.3 for USA.
2. *η* = 876, 824 (0.268% of population), the number of daily vaccinated individuals is estimated as the average of daily numbers of 30 days (February 23, 2021 to March 24, 2021) from the ourworldindata website (https://ourworldindata.org/).
3. *V*_0_ = 19, 882, 544, the number of initial fully-vaccinated individuals on February 23, 2021, is adopted from the ourworldindata website (https://ourworldindata.org/).
4. *k*_2_ = 1.82%, the ratio of dead and recovered individuals among confirmed cases, *C*(0) = 28, 302, 207, the initial confirmed cases, *D*_*c*_(0) = 503, 937, the initial confirmed deaths, and *A*(0) = 9, 196, 400 are adopted from Johns Hopkins University Coronavirus Center (*22*).
5. *R*_*C*_(0) = 28, 185, 069, the number of initial confirmed recovered individuals that still remain immunity to the virus is estimated with the recurrence formula *R*_*C*_(*t*) = (*C*(*t*) *− R*_*C*_(*t −* 1))*/t*_*s*_. *R*_*Q*_(0) is calculated with *R*_*Q*_(0) = *k*_1_*R*_*C*_(0).

Fig. 3 presents the prediction and inference of the epidemic in USA assuming a 8-month duration of immunity. The SUVQC model is fitted to the daily numbers of confirmed cases from February 23, 2021 to March 24, 2021, and the unknown parameters and variables are inferred to be *U*_*s*_(0) = 1, 590, 637, *Q*(0) = 747, 060, *γ*_1_ = 0.1603 and *β* = 0.1230 (Fig. 3A, red points, *R*^2^ = 0.9997). Suppose that all the inferred parameters remain constant values in future and the level of NPIs is retained similar to that of between February 23, 2021 and March 24, 2021 (the mean of the Government Stringency Index is 63.50 by OxCGRT. In comparison, the maximum Government Stringency Index of US is 75.46 in late November, 2020, and 65.91 between March 1, 2020 and March 24, 2021). The ending time of the epidemic of USA is estimated to be September 1, 2022, with 34, 672, 274 confirmed infected cases and 782, 183 confirmed deaths (Fig. 3, A and B). However, if we assume no NPIs with other parameters remain the same, the epidemic will end much later at February 28, 2027, with 62, 782, 387 confirmed infected cases and 1, 284, 642 confirmed deaths (Fig. 3, C and D). Notably that given a limit duration of immunity of 3-6 months, and the speed of vaccination, there is a probability that the epidemic never end without NPIs (Table 2, see the corresponding plots in Fig. 3, 5, and fig. S6, S7).

**Table 2:** Prediction of the epidemic in USA with different durations of immunity

| Duration of immunity | 90 days | 180 days | 240 days | 365 days |
| --- | --- | --- | --- | --- |
| **with NPIs** |  |  |  |  |
| Ending time | - | 2029-8-17 | 2022-9-1 | 2022-2-3 |
| Infected number | - | 43,635,623 | 34,672,274 | 32,908,322 |
| Death number | - | 942,400 | 782,183 | 750,653 |
| Maximum active number | 9,196,400 | 9,196,400 | 9,196,400 | 9,196,400 |
| Date of maximum active number | 2021-2-23 | 2021-2-23 | 2021-2-23 | 2021-2-23 |
| Equilibrium value of active number | 2,276,581 | 0 | 0 | 0 |
| **without NPI after Mar 24, 2021** |  |  |  |  |
| Ending time | - | - | 2027-2-28 | 2022-7-21 |
| Infected number | - | - | 62,782,387 | 44,675,741 |
| Death number | - | - | 1,284,642 | 960,991 |
| Maximum active number | 8,842,884 | 4,037,235 | 2,715,289 | 1,785,177 |
| Date of maximum active number | 2021-6-11 | 2021-6-25 | 2021-6-26 | 2021-3-24 |
| Equilibrium value of active number | 4,453,576 | 1,213,567 | 0 | 0 |

**Figure 3:**
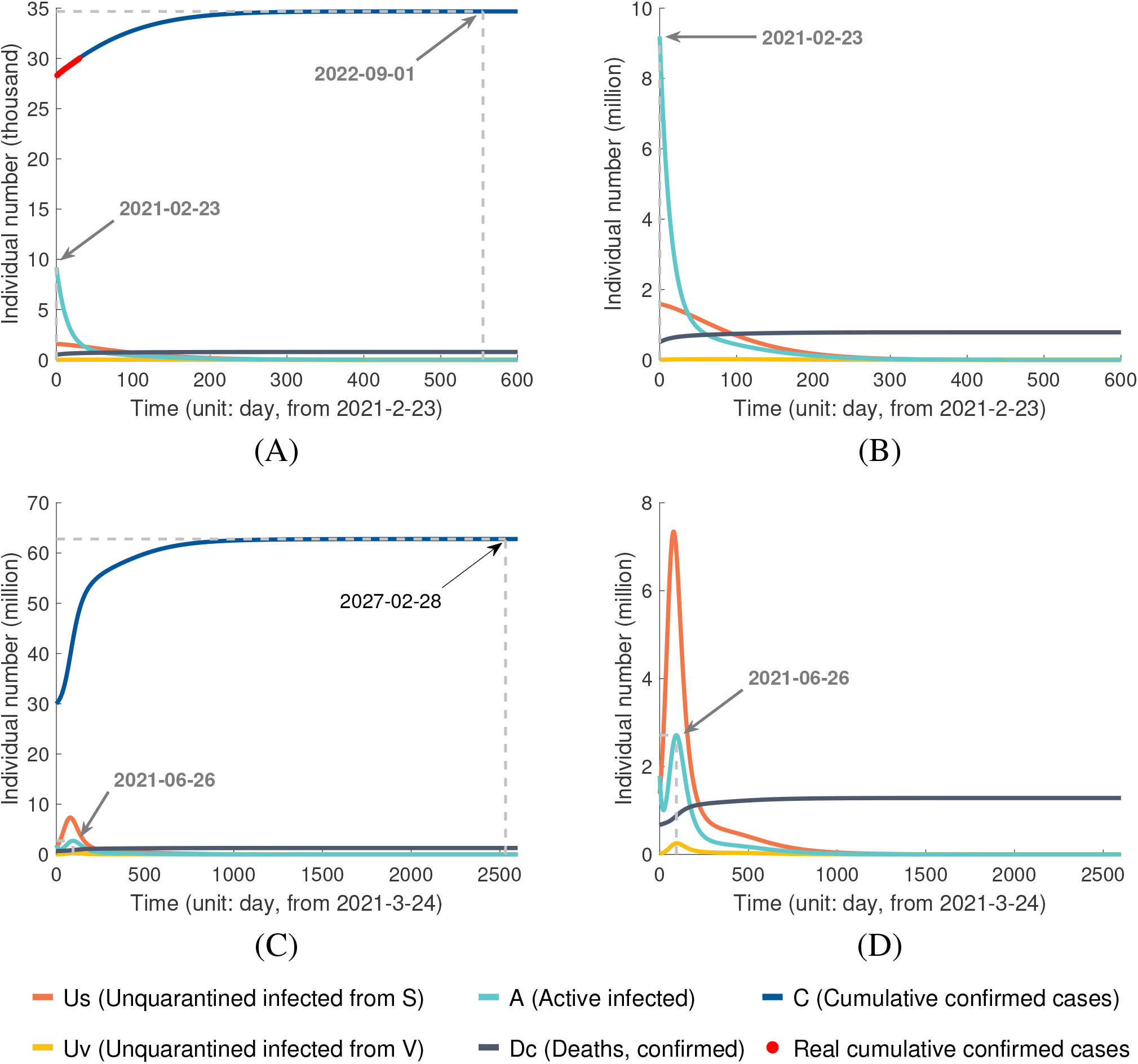
Prediction of epidemic dynamics in US with vaccination and multiple parameter settings: (A) and (B) Vaccine efficacy 1 *−λ*_1_ = 0.95, daily vaccination rate *η/N* = 0.268%, duration of immunity *t*_*s*_ = 8 months, and with NPIs. (C) and (D). All the other parameters are identical to (A) and (B), and with no NPIs.

**Figure 4:**
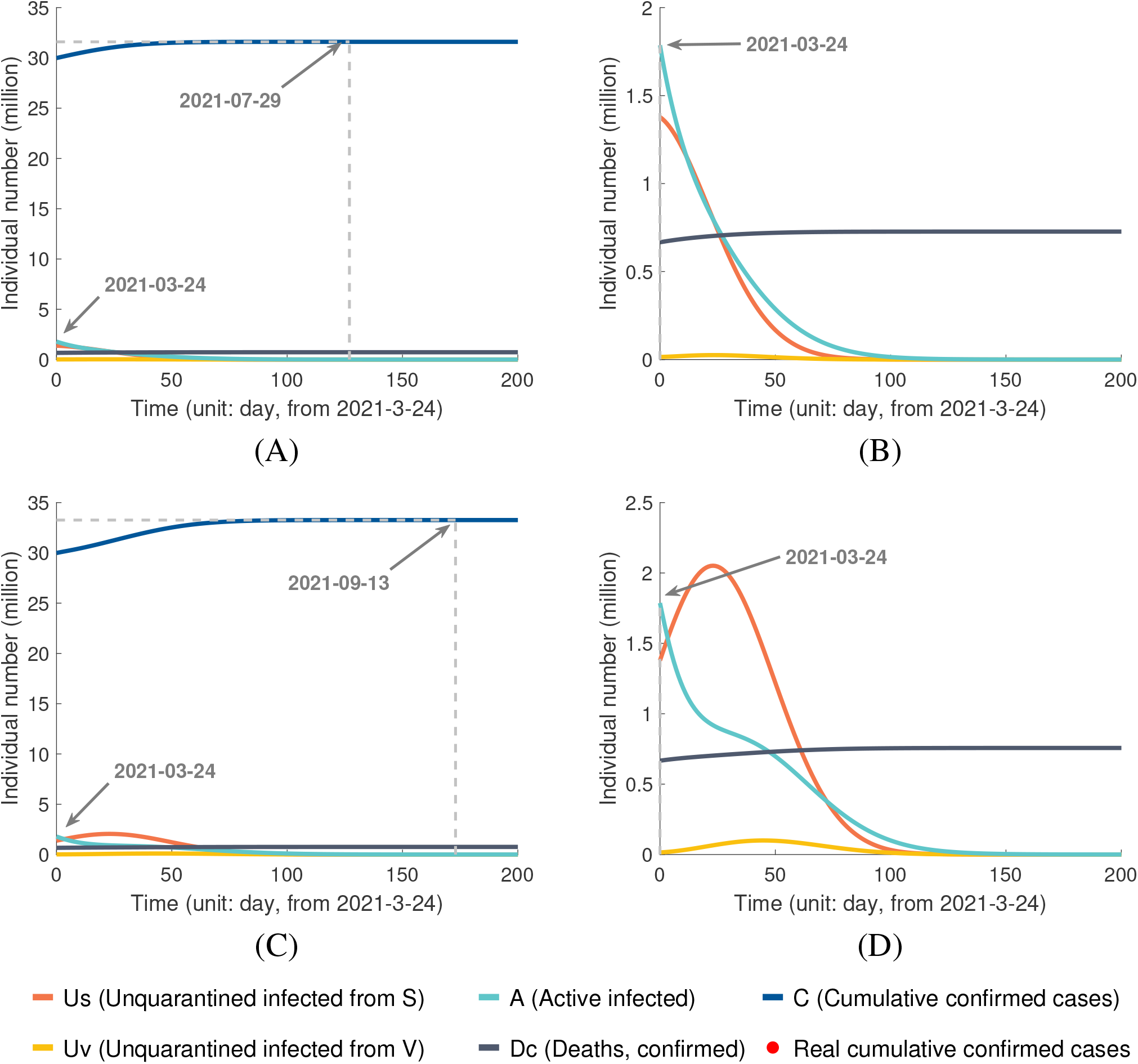
Prediction of epidemic dynamics in US with vaccination supposing a larger daily vaccination rate *η/N* = 1%: (A) and (B) Vaccine efficacy 1 *−λ*_1_ = 0.95, duration of immunity of 8 months, and with NPIs. (C) and (D). All the other parameters are identical to (A) and (B), and with no NPIs.

**Figure 5:**
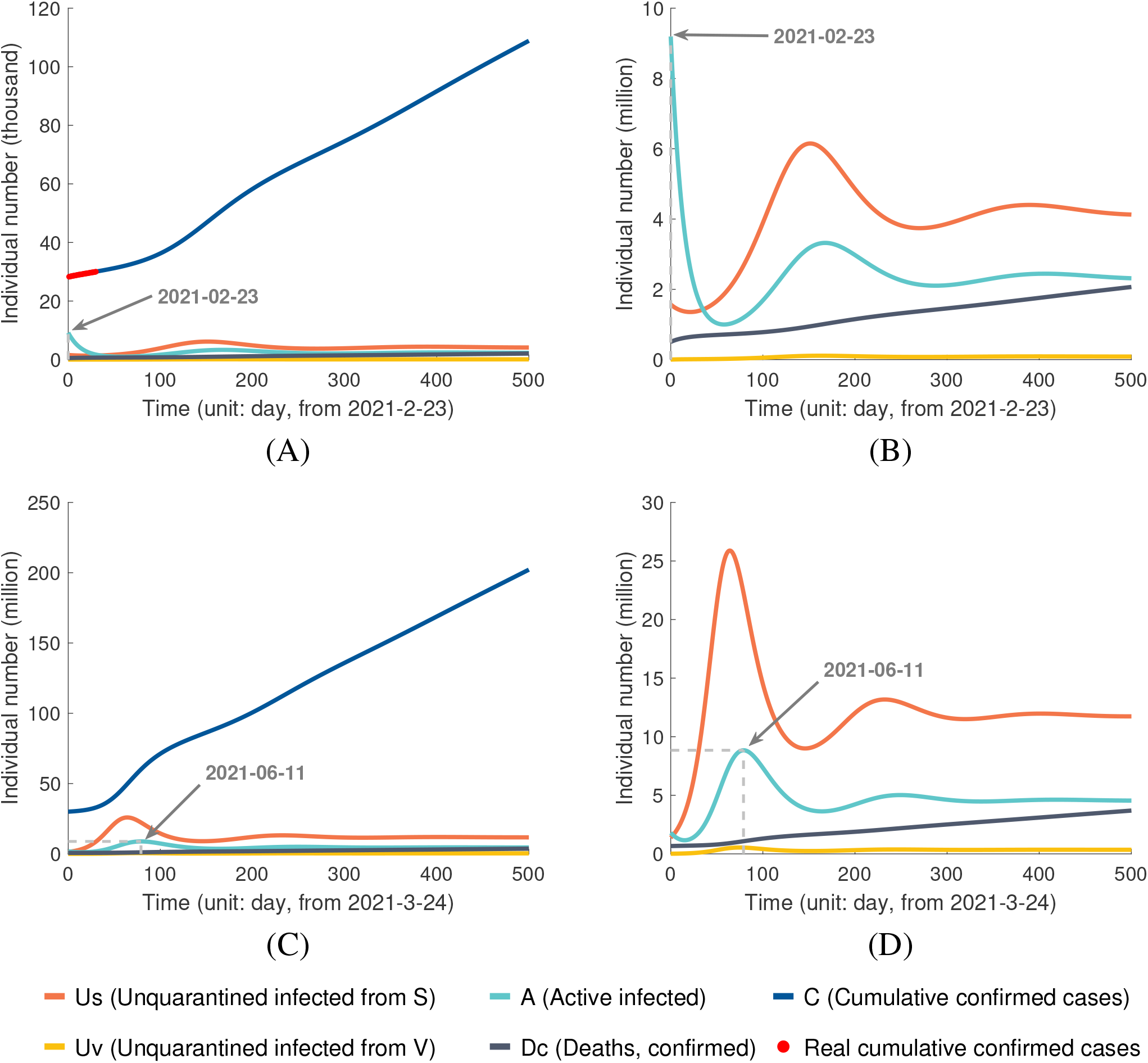
Prediction of epidemic dynamics in US under duration of immunization of three months: (A) and (B) Vaccine efficacy 1 *−λ*_1_ = 0.95, daily vaccination rate *η/N* = 0.268%, and with NPIs. (C) and (D). All the other parameters are identical to (A) and (B), and with no NPIs.

We thus explore the epidemic dynamics in USA under other scenarios with different immunity durations and a higher speed of vaccination (Table 3). With the accelerated vaccination rate of 1%*N* per day (around 4 times of the current vaccination rate) and with NPIs, the ending time of the epidemic in USA ranges between July 23, 2021 and September 24, 2021, corresponding to different immunity durations of 3, 6, 8 and 12 months (see the corresponding plots in Fig. 3, and fig. S8-S10). The number of cumulative confirmed infections is estimated to be between 31, 442, 008 and 32, 702, 833, and the number of deaths between 724, 434 and 746, 979. The epidemic in USA will last for much longer time if no NPIs after Mar 24, 2021, ranging between September 2, 2021 and January 20, 2022, with the number of cumulative confirmed infections between 32, 575, 585 and 43, 588, 222, and the number of deaths between 744, 704 and 941, 552. Significantly, assuming an immunity duration of three months and the current daily vaccination rate of *η* = 0.268%*N* per day, the epidemic in USA will never end (Table 2 and Fig. 5). The epidemic reaches an equilibrium state with 2, 276, 581 active infected cases if under the current intensity of NPIs, and an equilibrium state of 4, 453, 576 active infected cases without NPI.

**Table 3:** Prediction of the epidemic in USA with different durations of immunity and accelerated vaccination of 1% N per day

| Duration of immunity | 90 days | 180 days | 240 days | 365 days |
| --- | --- | --- | --- | --- |
| **with NPIs** |  |  |  |  |
| Ending time | 2021-9-24 | 2021-8-6 | 2021-7-29 | 2021-7-23 |
| Infected number | 32,702,833 | 31,767,383 | 31,597,332 | 31,442,008 |
| Death number | 746,979 | 730,252 | 727,211 | 724,434 |
| Maximum active number | 1,792,512 | 1,787,972 | 1,786,647 | 1,785,177 |
| Date of maximum active number | 2021-3-24 | 2021-3-24 | 2021-3-24 | 2021-3-24 |
| Equilibrium value of active number | 0 | 0 | 0 | 0 |
| **without NPI after Mar 24, 2021** |  |  |  |  |
| Ending time | 2022-1-20 | 2021-9-25 | 2021-9-13 | 2021-9-2 |
| Infected number | 43,588,222 | 34,194,285 | 33,261,982 | 32,575,585 |
| Death number | 941,552 | 773,638 | 756,973 | 744,704 |
| Maximum active number | 2146,505 | 1,787,972 | 1,786,647 | 1,785,177 |
| Date of maximum active number | 2021-5-28 | 2021-3-24 | 2021-3-24 | 2021-3-24 |
| Equilibrium value of active number | 0 | 0 | 0 | 0 |

## Formula for optimal design of vaccination to achieve herd immunity

Let *f* be the proportion of the population that retains immunological memory by vaccination at time *t, f* = *V/N*. We consider a discrete transmission model for simplicity, with *t*_*i*_ denote the *i*-th transmission cycle. Let *M* (*t*_*i*_) denote the number of new infected cases at time *t*_*i*_, it includes two parts: the un-quarantined infected individuals from vaccinated individuals, *U*_*v*_(*t*_*i*_) = *M* (*t*_*i*_)*fλ*_1_*/*(*fλ*_1_ + (1 *− f*)), and from susceptible individuals *U*_*s*_(*t*_*i*_) = *M* (*t*_*i*_)(1 *− f*)*/*(*fλ*_1_ + (1 *− f*)). The number of new infected cases at the next time step *t*_*i*+1_ is

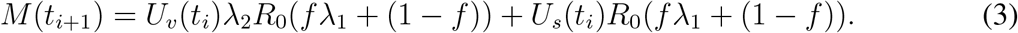

By definition the effective reproductive number of time *t*_*i*_ can be calculated with

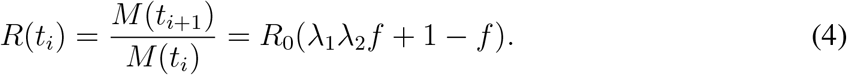

For an effective control of the epidemic, we should have *R*(*t*_*i*_) *<* 1, which is

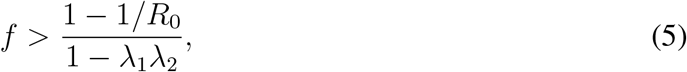

and it is straightforward to get the speed of vaccination

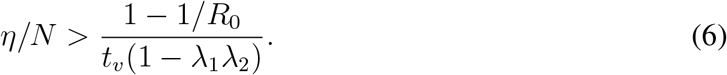

Note when *V* (*t*_*i*_) = *N*, the maximum value of *η*(*t*_*i*+1_)*/N* = 1*/t*_*v*_, which implies that the long-term upper bound of *η/N* is 1*/t*_*v*_. Thus from Eqn 6, we conclude that when 1 *−* 1*/R*_0_ *>* 1 *− λ*_1_*λ*_2_, the epidemic can not be terminated by vaccination solely. For example, assume that the basic reproductive number of SARS-CoV-2 is *R*_0_ = 2.5, and that *U*_*v*_ has the same infectivity as *U*_*s*_ (that is, *λ*_2_ = 1), *λ*_1_ should be smaller than 0.4 (correspondingly, the efficacy of vaccine is *≥* 0.6) to ensure ending the epidemic by vaccination only.

Equation 6 provides a useful formula for choosing vaccination rate taking into account of the different values of *R*_0_, *t*_*v*_ and *λ*_1_ to guarantee ending the epidemic. For *R*_0_ = 2.5, *t*_*v*_ = 100 days, *λ*_1_ = 0.4 (e.g., the efficacy of the vaccines 0.6), *λ*_2_ = 1, *η* should be *>* 1% of the population per day. For *R*_0_ = 2.5, *t*_*v*_ = 100 days, *λ*_1_ = 0.05 (e.g., the efficacy of the vaccines 0.95), *λ*_2_ = 1, *η* should *>* 0.63% of the population per day (Fig. 6, red point). Fig. 6 provides a thorough demonstration of the minimum vaccination rates to assure effectively ending the epidemic for a wide range of *R*_0_ and vaccine efficacy values.

**Figure 6:**
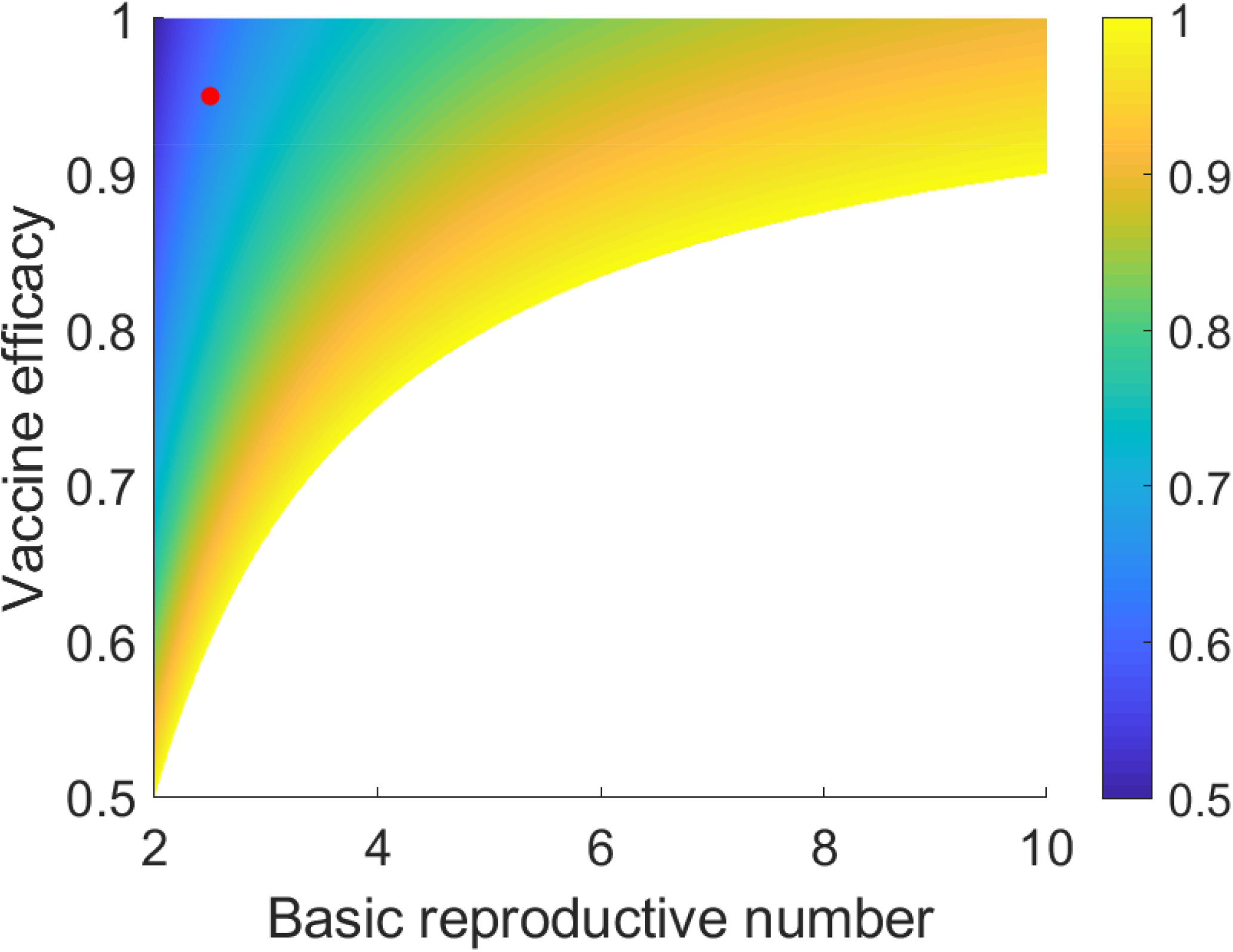
The minimum daily vaccination rate *η/N* required to effectively end the epidemic without NPIs for different values of basic reproductive number (*R*_0_) and vaccine vaccine efficacy (1*− λ*_1_). Here we set *λ*_2_ = 1. The color scheme indicates the daily vaccination rates in unit of 1*/t*_*v*_. Red dot: for *R*_0_ = 2.5, *t*_*v*_ = 100 days and 1*− λ*_1_ = 0.95, the minimum daily vaccination rate should be *η/N >* 0.63% per day.

## Discussion

In this study, we develop a model SUVQC to characterize the pandemic dynamics of COVID-19 under vaccination and NPIs. In addition to the intrinsical parameters of the epidemic, such as transmissibility, and the effect of NPIs, the model also explicitly parameterizes multiple key factors related to vaccination, including the vaccine efficacy, the vaccination rate, and the reduced infectivity of individuals who have been vaccinated and infected etc.

Note that SUVQC has some strong assumptions, which may bias the inference and prediction. The model assumes that all individuals are equally susceptible in the epidemic. In reality, individuals in the hot spots of human interaction network are in a higher exposure risk level. A vaccination program prioritizing these individuals are more effective to contain the epidemic, especially when there is a shortage of vaccines. For such a vaccination program, the daily vaccination rate provided by Eqn 6 can be overestimated. Furthermore, the model includes 12 parameters, for most of which the data (daily reported number of confirmed cases) is uninformative, and has to be inferred beforehand with prior knowledge. Some critical parameters, such as the duration of effective immunity, is still unclear with limited studies. The prediction of epidemic by the model are made based on some pre-selected values.

The model is applied to the numbers of daily reported confirmed cases to project the future trends of Israel and USA with the parameters inferred based on their current epidemic status and intervention measures. The analysis demonstrates that a high vaccine efficacy and a high vaccination rate coupled with intensive NPIs can eliminate the spread of the virus very efficiently. Although for USA, under some parameter settings, such as, a short duration of immunity, or with no NPIs, the epidemic will not end and instead reach an equilibrium state with a proportion of the population remaining actively infected. We further provide a unified formula for determining the minimum daily vaccination rate for different values of effective reproductive number and vaccine efficacy. Considering the vaccine parameters and epidemic parameters are difficult to adjust in operation, a practical scheme is to supplement with NPIs when implementing the vaccination plan.

However, there are many reasons to be conservative about the efficacy of vaccination in suppressing the epidemics. The SARS-CoV-2 genomes are still actively evolving under selection with new mutations, and SARS-CoV-2 may gain function to partially escape the vaccine-induced immunity and reduce the vaccine efficacy (*24–28*). It is possible that current vaccines may have reduced efficacy and immunity duration in the near future. In addition, many COVID-19 reinfection reports imply the immunity may not so strong and long-lasting (*29,30*). Excessive optimism is undesirable considering the unpredictable factors. Retaining NPIs at least in some extend is still necessary to contain the pandemic. If the vaccination program works, the pandemic in Israel should end by mid-May (no later than August 26, 2021, assuming there is no NPIs and the duration of immunity is 180 days). If the does not end by then, we must re-evaluate the role of vaccination.

## Data Availability

only public data were used.

## Acknowledgments

This study was supported by the National Key R&D Program of China (Grant No. 2020YFC0847000) and the National Natural Science Foundation of China (Grant Nos. 31571370, 91731302, and 31772435).

## Supplementary Materials

**Figure S.1:**
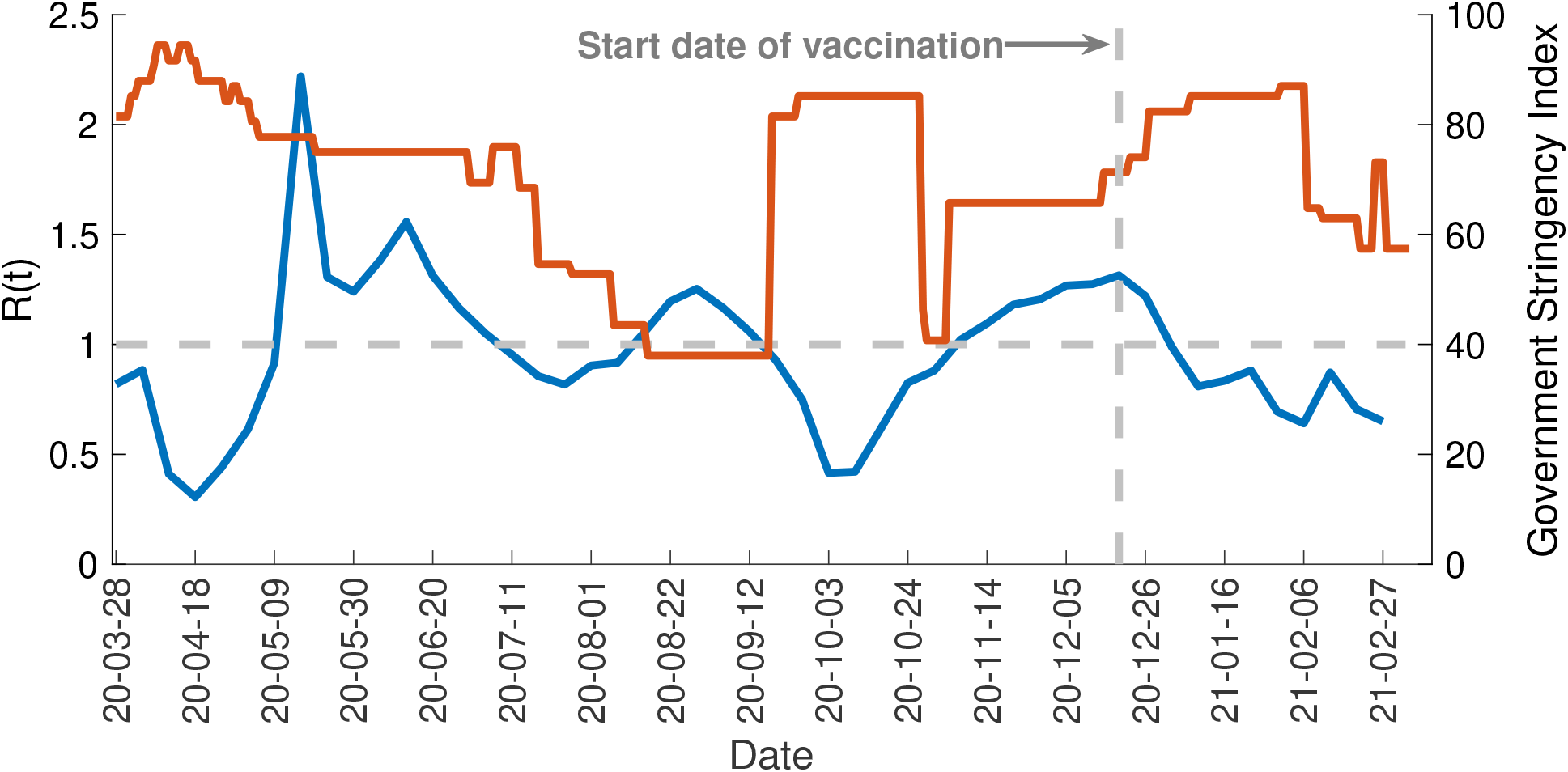
Temporal dynamics of R(t) and the Government Stringency Index in Israel. Blue line indicates R(t) value estimated with the SUQC model. Ted line is the Government Stringency Index adopted from OxCGRT.

**Figure S.2:**
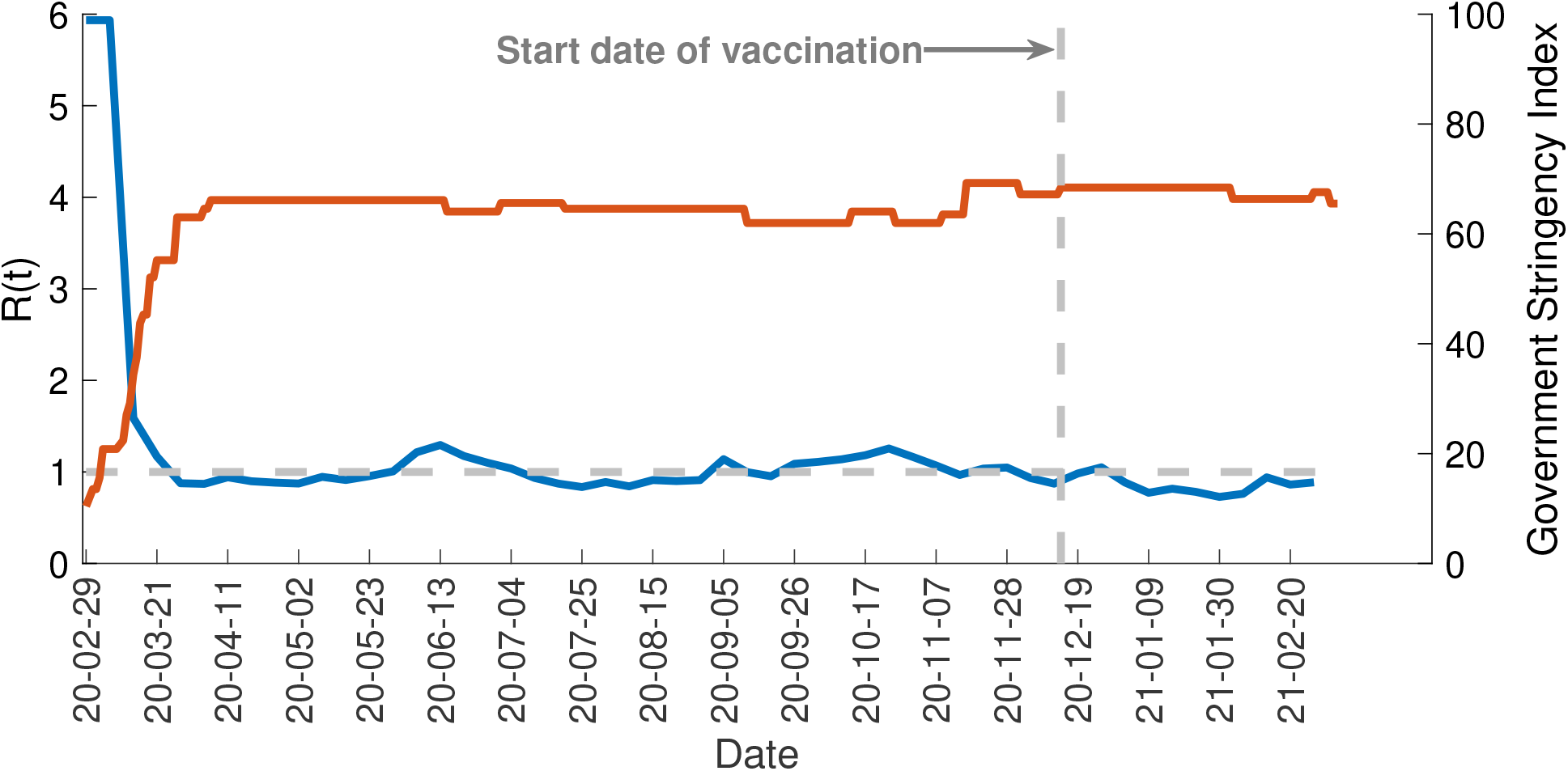
Temporal dynamics of R(t) and the Government Stringency Index in US. Blue line indicates R(t) values estimated with the SUQC model. Ted line is the Government Stringency Index adopted from OxCGRT.

**Figure S.3:**
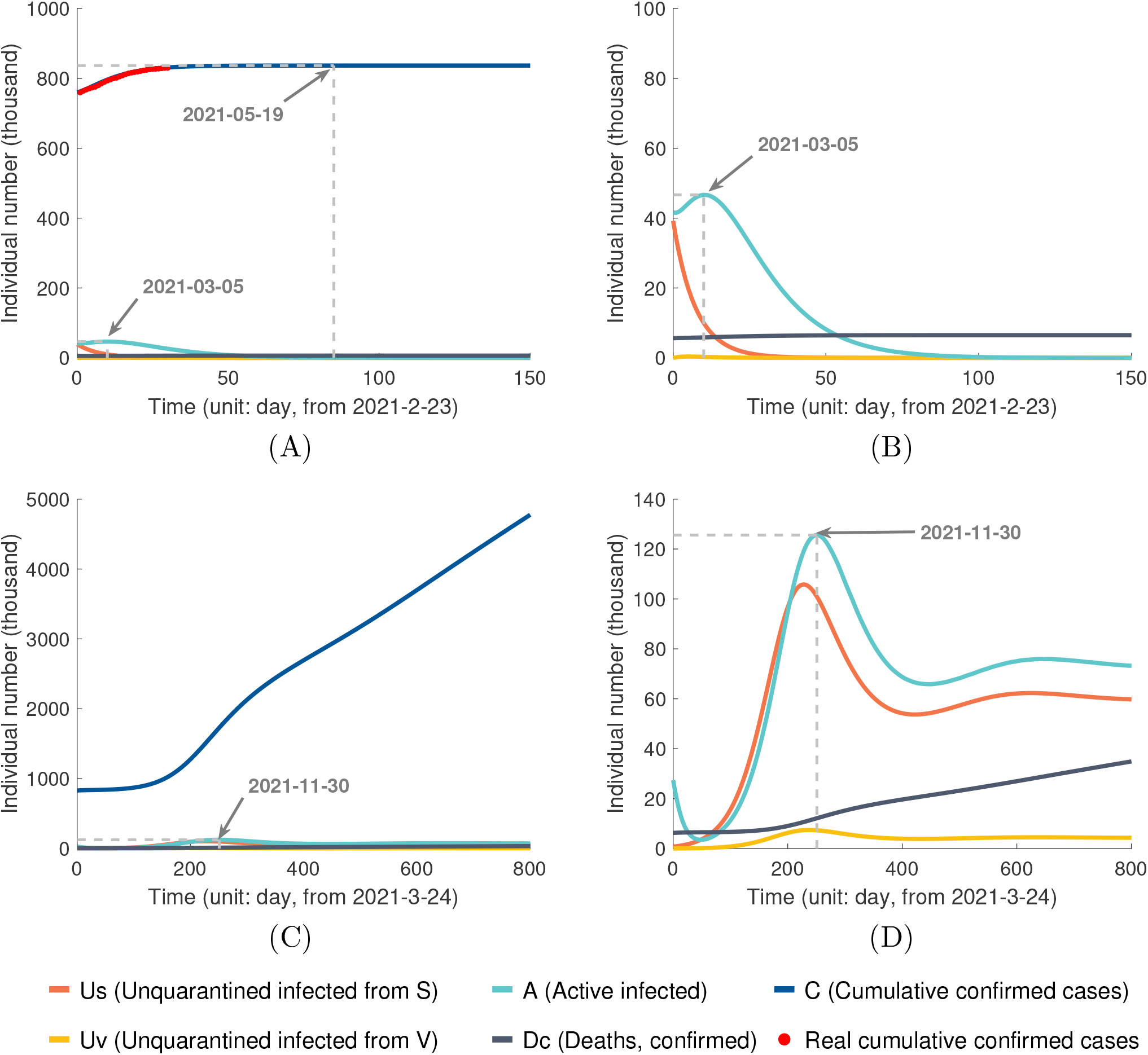
Prediction of epidemic dynamics in Israel with the immunity duration of three months: (A) and (B) Vaccine efficacy 1 *− λ*_1_ = 0.95, daily vaccination rate *η/N* = 0.605%, and with NPIs. (C) and (D). All the other parameters are identical to (A) and (B), and with no NPIs.

**Figure S.4:**
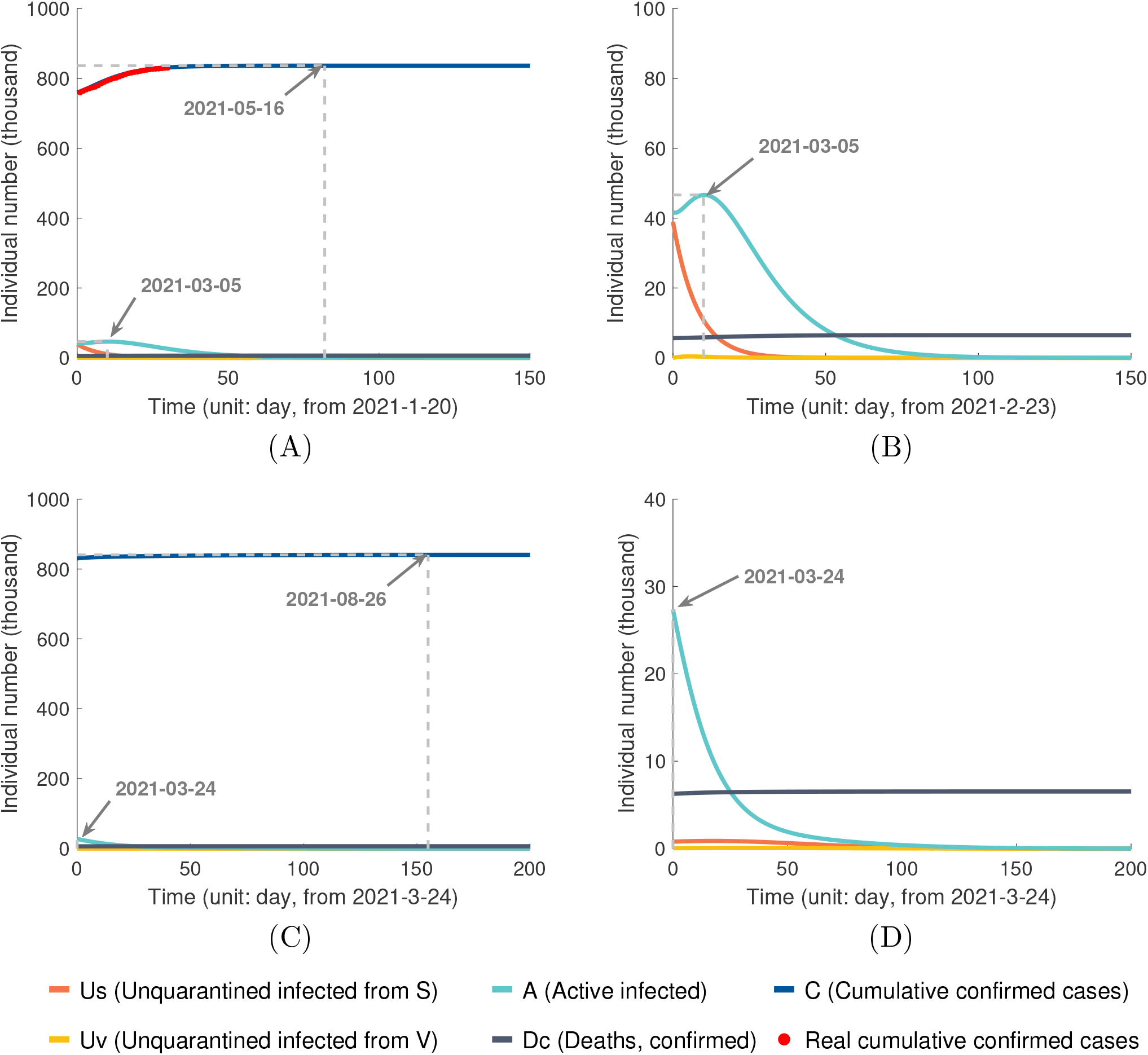
Prediction of epidemic dynamics in Israel with the immunity duration of six months: (A) and (B) Vaccine efficacy 1 *− λ*_1_ = 0.95, daily vaccination rate *η/N* = 0.605%, and with NPIs. (C) and (D). All the other parameters are identical to (A) and (B), and with no NPIs.

**Figure S.5:**
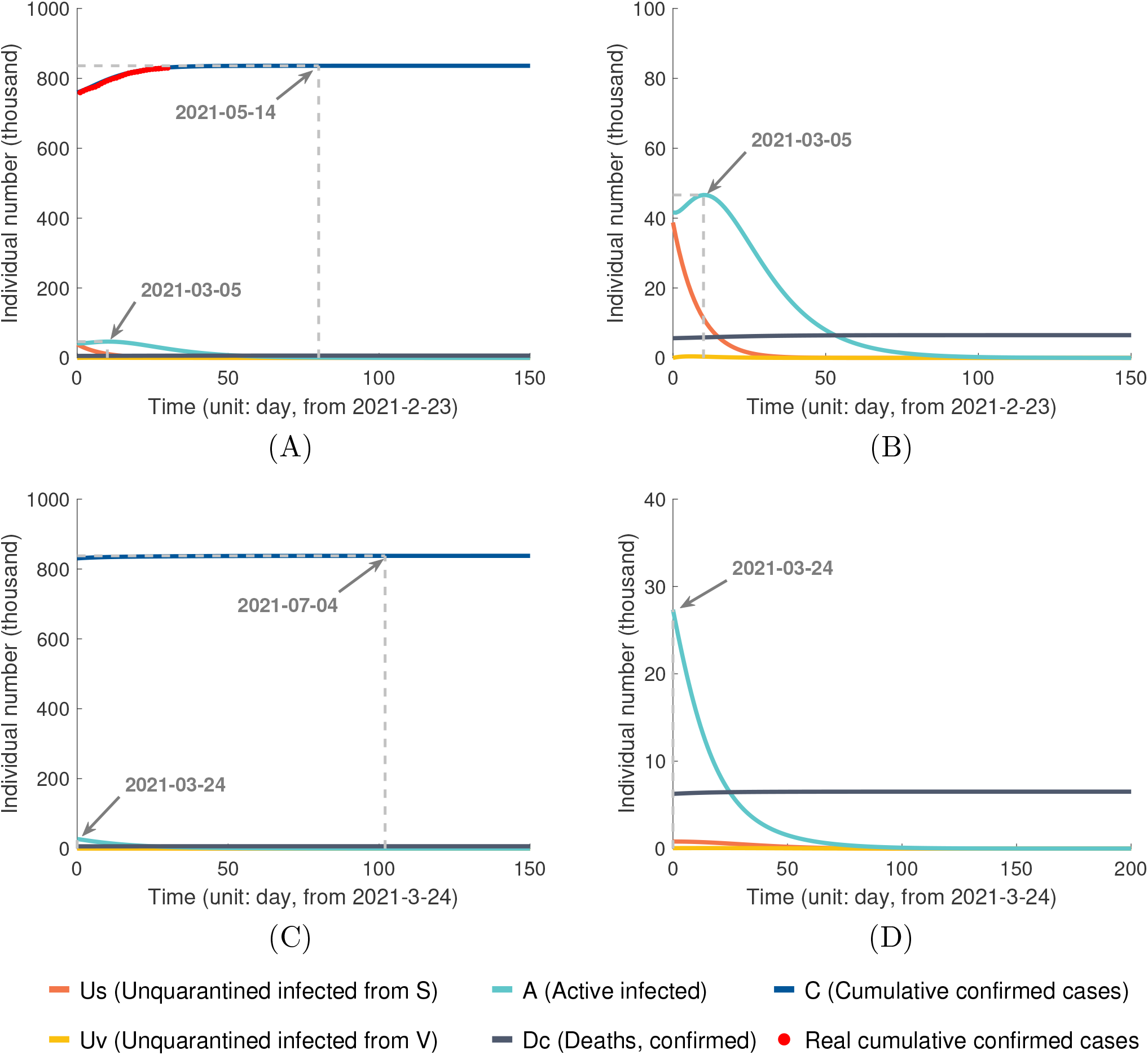
Prediction of epidemic dynamics in Israel with the immunity duration 12 months: (A) and (B) Vaccine efficacy 1 *− λ*_1_ = 0.95, daily vaccination rate *η/N* = 0.605%, and with NPIs. (C) and (D). All the other parameters are identical to (A) and (B), and with no NPIs.

**Figure S.6:**
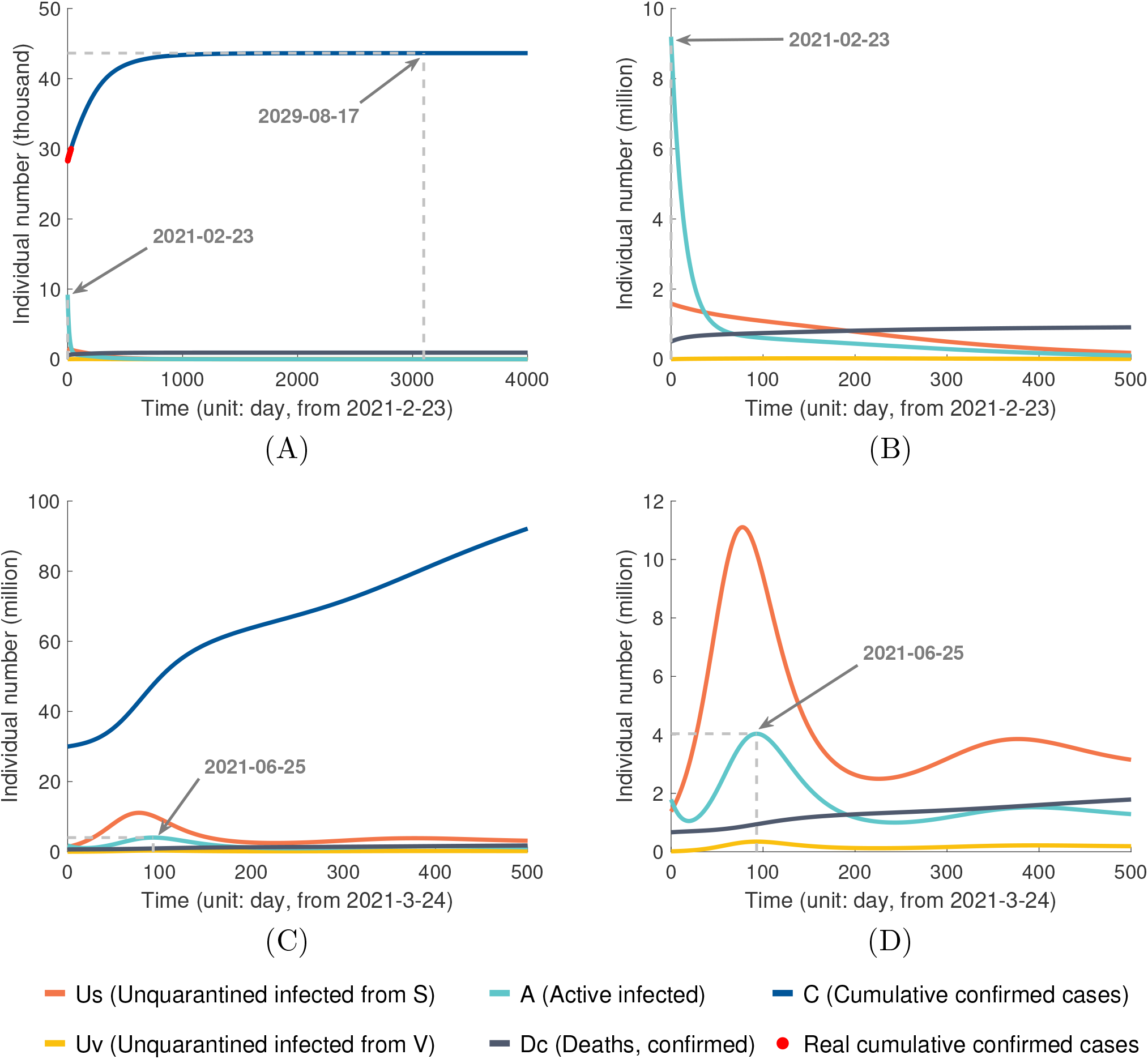
Prediction of epidemic dynamics in USA with the immunity duration of six months: (A) and (B) Vaccine efficacy 1 *− λ*_1_ = 0.95, daily vaccination rate *η/N* = 0.268%, and with NPIs. (C) and (D). All the other parameters are identical to (A) and (B), and with no NPIs.

**Figure S.7:**
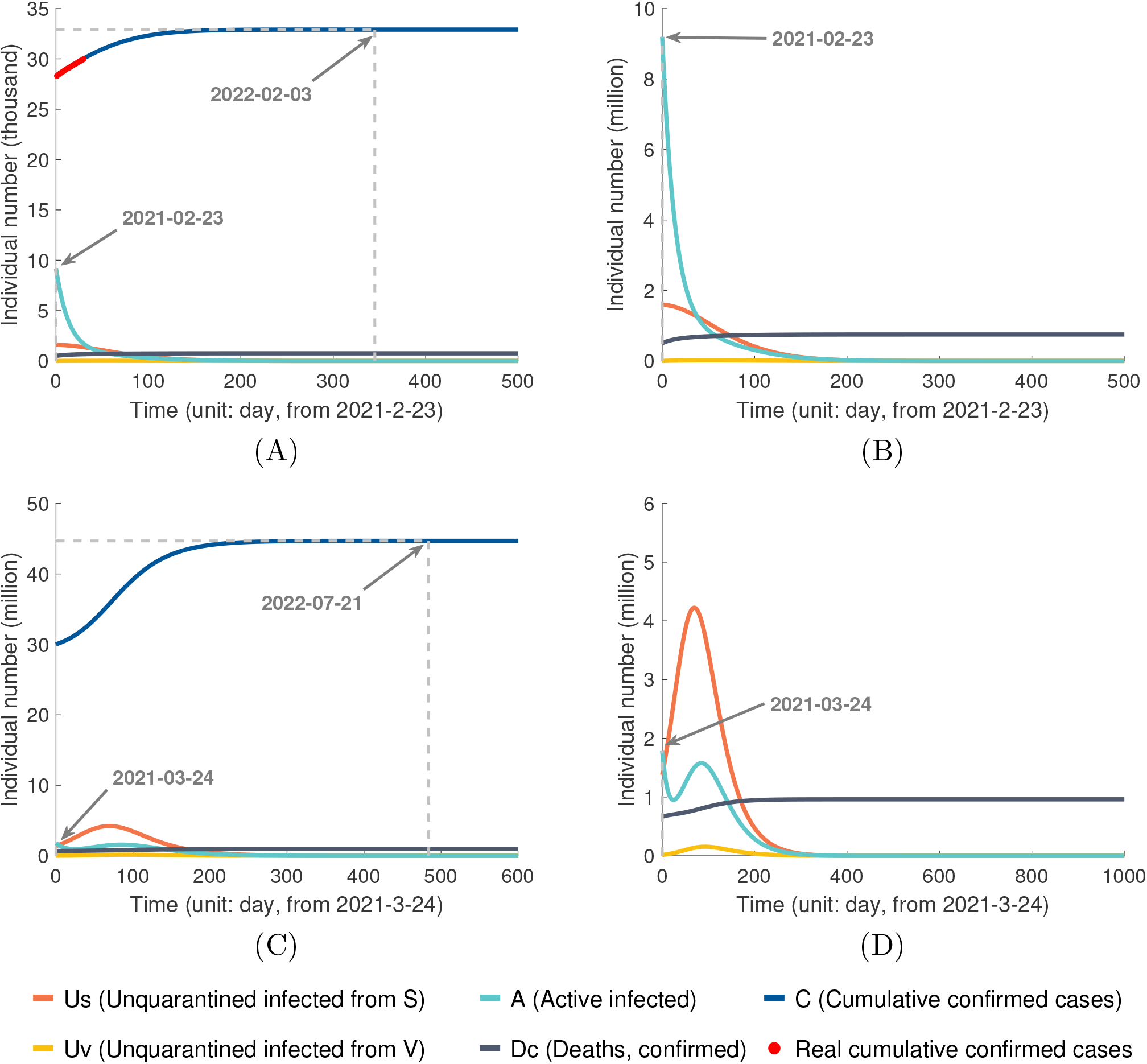
Prediction of epidemic dynamics in USA with the immunity duration of 12 months: (A) and (B) Vaccine efficacy 1 *− λ*_1_ = 0.95, daily vaccination rate *η/N* = 0.268%, and with NPIs. (C) and (D). All the other parameters are identical to (A) and (B), and with no NPIs.

**Figure S.8:**
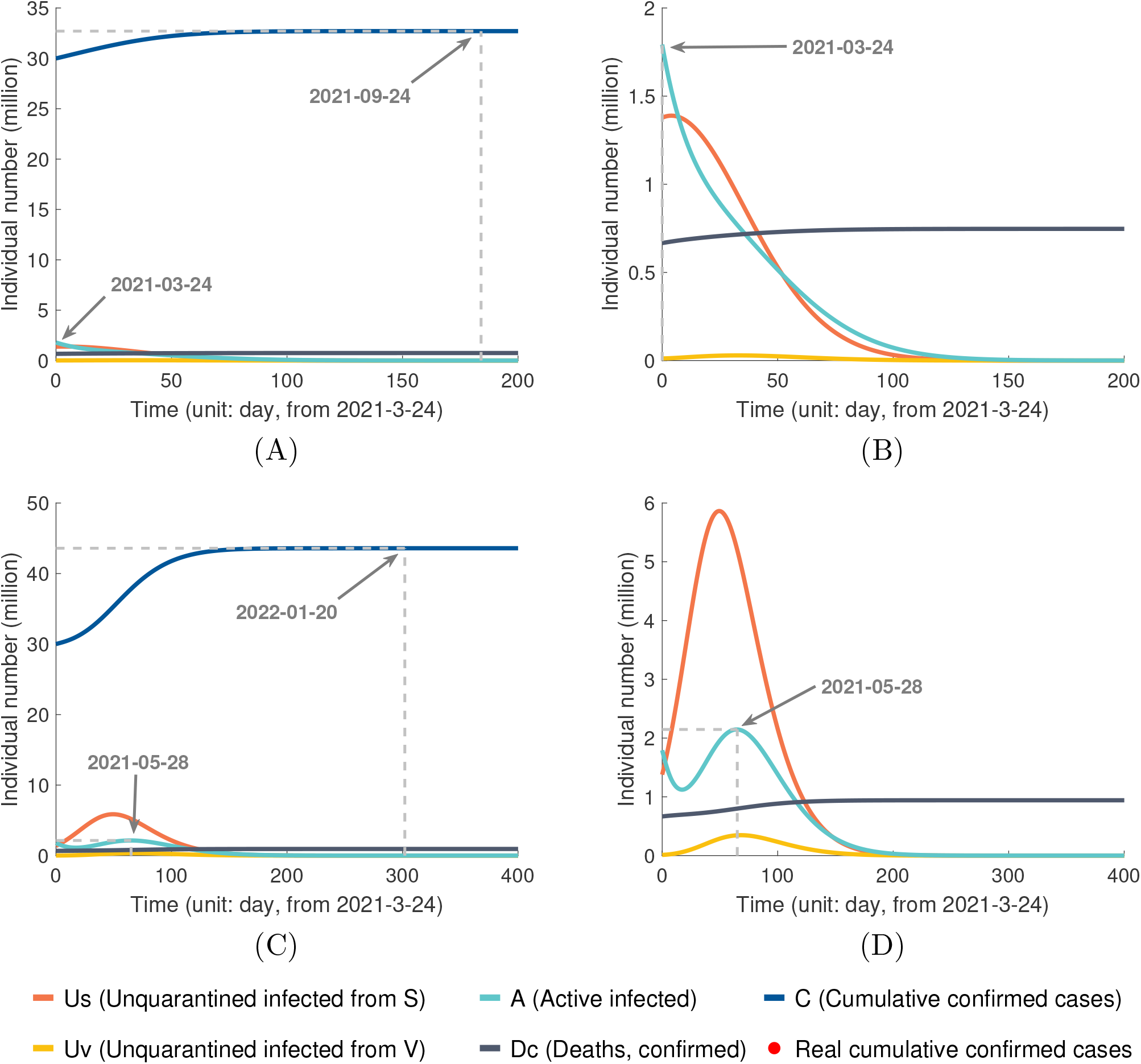
Prediction of epidemic dynamics in USA with the immunity duration of three months and daily vaccination rate *η* = 0.01*N* : (A) and (B) Vaccine efficacy 1 *− λ*_1_ = 0.95 and with NPIs. (C) and (D). All the other parameters are identical to (A) and (B), and with no NPIs.

**Figure S.9:**
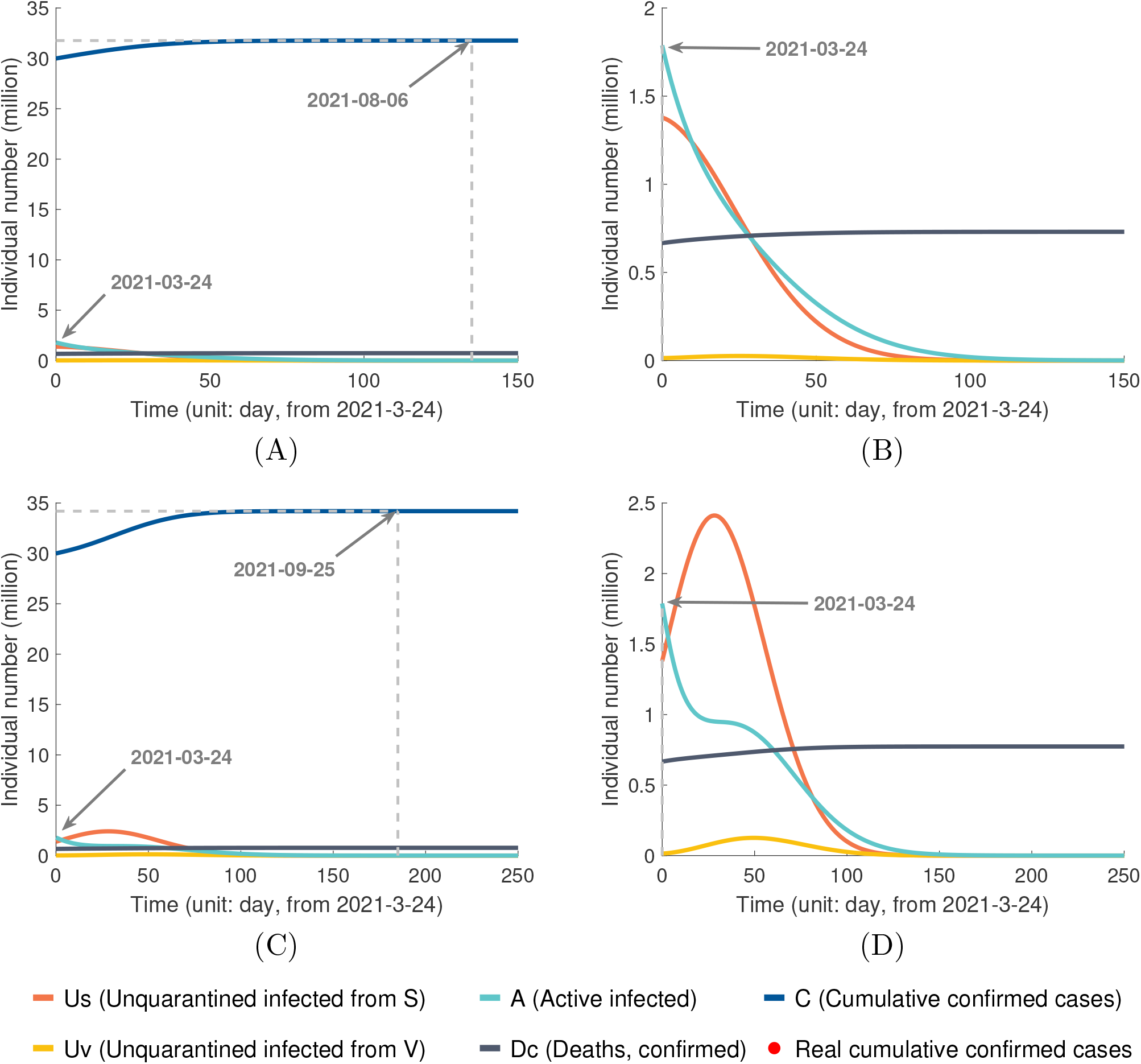
Prediction of epidemic dynamics in USA under duration of immunization of six months and daily vaccination rate *η* = 0.01*N* : (A) and (B) Vaccine efficacy 1 *− λ*_1_ = 0.95 and with NPIs. (C) and (D). All the other parameters are identical to (A) and (B), and with no NPIs.

**Figure S.10:**
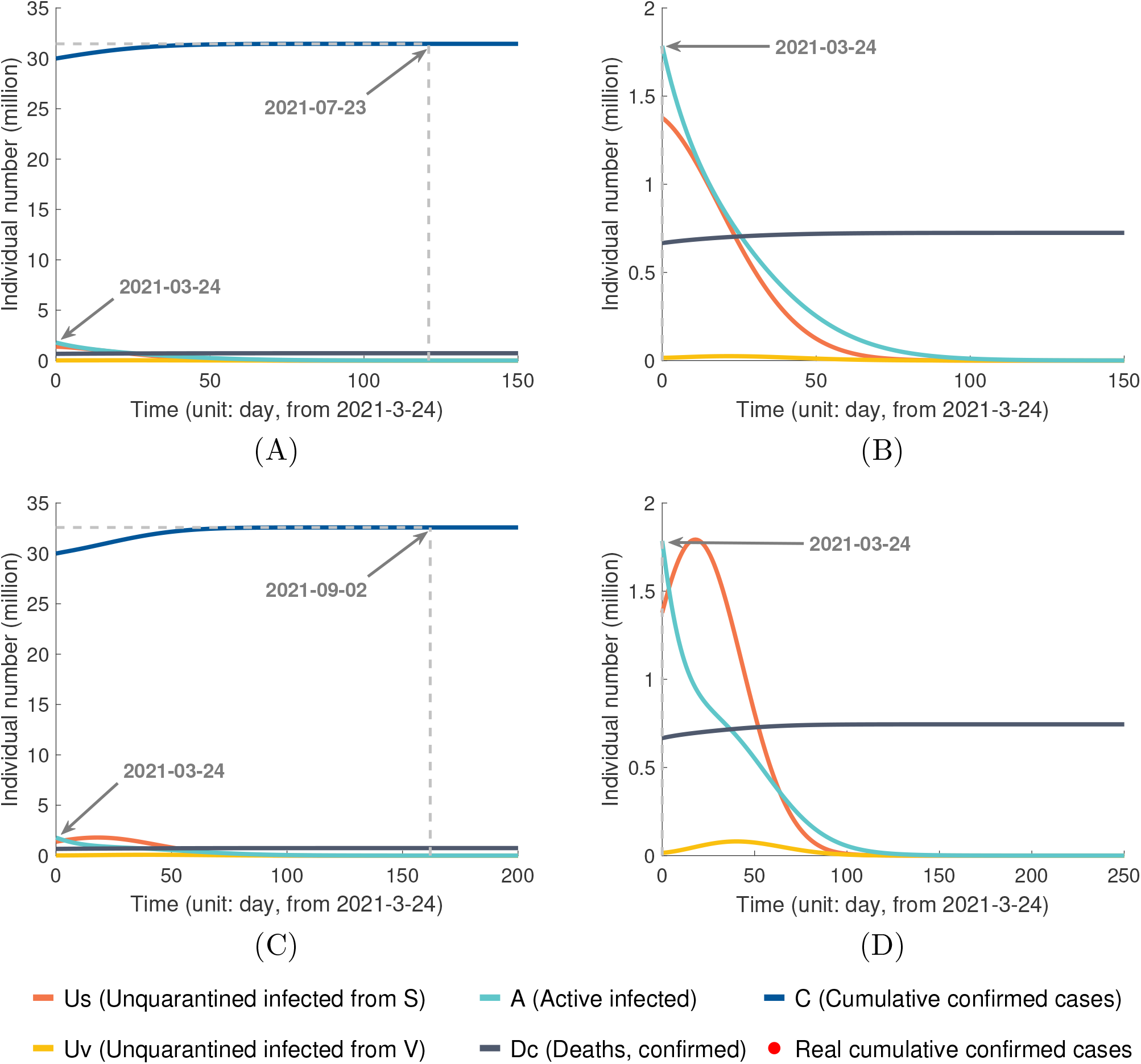
Prediction of epidemic dynamics in US under duration of immunization of 12 months and daily vaccination rate *η* = 0.01*N* : (A) and (B) Vaccine efficacy 1 *− λ*_1_ = 0.95 and with NPIs. (C) and (D). All the other parameters are identical to (A) and (B), and with no NPIs.

